# Prevalence of Mental Health Disorders among Elderly Diabetics and Associated Risk Factors in Indonesia

**DOI:** 10.1101/2020.11.25.20238659

**Authors:** Mahalul Azam, Rina Sulistiana, Arulita Ika Fibriana, Soesmeyka Savitri, Syed Mohamed Al Junid

## Abstract

This cross-sectional study aimed to explore the prevalence of mental health disorders (MHD) among elderly diabetics in Indonesia and their associated risk factors. Data were extracted from the 2018 national basic health survey, Indonesia (abbreviated as the acronym of RISKESDAS). The survey involved households randomly selected from 34 provinces, 416 districts, and 98 cities in Indonesia, with 1,017,290 respondents. The number of subjects selected in this study was 2,818 elderly diabetic subjects. MHD was determined by self-reporting assessment. Secondary data acquired from the RISKESDAS 2018 data involved age, sex, urban-rural residence status, marital status, educational level, employment status, obesity, hypertension, heart disease, stroke, family history of MHD, and duration of DM. Binary logistic regression was used to analyze the risk factors related to MHD among elderly diabetics. Prevalence of MHD among elderly diabetics in Indonesia was 19.3%. Factors associated with MHD among elderly diabetics were obesity (prevalence odds ratio [POR]=4.57; 95% CI: 3.312-6.297), family history of MHD (POR=2.43; 95% CI: 1.707-3.471), lower education (POR=1.93; 95% CI: 1.464-2.533), stroke (POR=1.76; 95% CI: 1.292-2.384), hypertension (POR=1.74; 95% CI: 1.416-2.145), heart diseases (POR=1.49; 95% CI: 1.123-1.973), female (POR=1.43; 95% CI: 1.122-1.813), and urban residence (POR=0.75; 95% CI: 0.607-1.183). The prevalence of MHD among elderly diabetics in Indonesia was 19.3%, suggesting that screening for psychological problems and educating elderly diabetic patients is essential. Obesity, family history of MHD, lower education, stroke, hypertension, heart disease, female, and rural residence altogether more likely to experience MHD in elderly diabetics.

## Introduction

In 2019, it was reported that 463 million individuals globally suffered from diabetes mellitus (DM). It was increased from 382 million in 2013[1]. The united States, China, India, and Indonesia are countries with a high prevalence of DM[2]. The prevalence of DM in Indonesia was 5.7% in 2007 and increased by 10.9% in 2018 [3, 4] caused a number of 157,500 or 6% of total deaths[5]. DM was a catastrophic and financial burden disease that expensed USD 381.25 million for all DM related hospital treatment in 2019 based on national health insurance (Jaminan Kesehatan Nasional=JKN)[6].

Mental health disorder (MHD) is the most frequent comorbid for DM with a prevalence of 28% globally; females tend to be higher than males, i.e., 34% and 23%, respectively[7–10]. Previous mental disorder conditions such as generalized anxiety disorder (GAD), major depressive disorder (MDD), bipolar disorder, and eating disorders are the underlying comorbid for DM patients with MHD[11–14] MHD in diabetics may decrease quality of life[15], poor self-care management[16], increase disability[17], cardiovascular mortality risk[18] and all-cause mortality risk[19]. On the other side, diabetes is a risk factor for MHD[20].

Younger diabetics are more likely to get MHD[8] while another study reported that elderly diabetics more likely to get MHD, with the increased risk of other factors present[11]. The previous study also concluded that MHD is more likely to occur in females, no formal education, current alcohol abusers, type I DM, longer duration of DM, chronic complication of DM, and other comorbidities among elderly diabetics patients[21]. However, there is a lack of information regarding MHD risk factors among elderly diabetics in Indonesia. The five-annual national basic health survey (abbreviated as an acronym of RISKESDAS: riset kesehatan dasar) 2018[3] was the latest national survey conducted by the Ministry of Health, Republic of Indonesia. The present study aimed to determine the prevalence and risk factors of MHD among elderly diabetics in Indonesia.

## Materials and Methods

### Design and study population

This cross-sectional study employed secondary data acquired from RISKESDAS 2018, which is the latest round of study. The survey involved households randomly selected from 34 provinces, 416 districts, and 98 cities in Indonesia, with 1,017,290 respondents[3]. The study population involved elderly diabetics in the survey aged older than 60 years old. Diabetics status was determined based on the questionnaire as well as age of the respondents. Diabetics status was defined as answering ‘Yes’ to the question delivered, whether subjects had diagnosed DM by the doctor. Subjects with incomplete data were excluded from the study. More details regarding data collection, ethical issues, and other related steps published in the RISKESDAS 2018 report[3].

### Data collection

This study was approved by the Ethics Committee, the National Institute of Health Research and Development (NIHRD), the Ministry of Health, Republic, Indonesia. MHD status was determined by the WHO self-reporting questionnaire-20 (SRQ-20),[22] as acquired from RISKESDAS 2018 data. Secondary data were also acquired from RISKESDAS 2018 that involved age, sex, urban-rural residence status, marital status, educational level, employment status, obesity, hypertension, heart disease, stroke, family history of MHD, and duration of DM.

### Statistical analysis

Subject’s characteristics were presented as frequency and proportions. The relationships of the determinants and MHD status were analyzed by the Chi-square test. The p-values <0.05 were considered statistically significant. Parameters that had p-value <0.25 then involved in the multivariate analysis using binary logistic regression. All statistical analyses were performed using the Statistical Package for the Social Sciences (SPSS) software (version 23.0 for Windows, IBM SPSS Inc., Chicago, IL).

## Results

Data extracted from RISKESDAS 2018 contained 2,818 elderly diabetics subjects. Table 1 showed that the sex proportion of elderly diabetics in the study population was higher in females, while the age category was almost comparable. Most of the elderly diabetics had lower education, live in the urban area, unemployed, and married. A few amounts of the study population, i.e., around 6%, were obese as well as with a family history of MHD. Hypertension and duration of DM were almost comparable, while heart disease and stroke had a lower proportion in the total population. The overall prevalence of MHD among elderly diabetics was 19.3% of the study population.

**Table 1.** Subject’s characteristics

| Characteristics | Frequency (n) | Percentage (%) |
| --- | --- | --- |
| **Sex** |  |  |
| Male | 1163 | 41.3% |
| Female | 1655 | 58.7% |
| **Age** |  |  |
| 60-69 years | 1489 | 52.8% |
| >69 years | 1329 | 47.2% |
| **Education level** |  |  |
| JHS or lower | 2049 | 72.7% |
| SHS or higher | 769 | 27.3% |
| **Residence** |  |  |
| Urban | 1799 | 63.8% |
| Rural | 1099 | 36.2% |
| **Employment status** |  |  |
| Unemployed | 1723 | 61.1% |
| Employed | 1095 | 38.9% |
| **Marital status** |  |  |
| Not married | 1182 | 41.9% |
| Married | 1636 | 48.1% |
| **Obesity** |  |  |
| Yes | 189 | 6.7% |
| No | 2629 | 93.3% |
| **Family history of MHD** |  |  |
| Yes | 171 | 6.1% |
| No | 2647 | 93.9% |
| **Duration of DM** |  |  |
| ≤ 5 years | 1555 | 55.2% |
| ≥ 5 years | 1263 | 44.8% |
| **Hypertension** |  |  |
| Yes | 1441 | 51.1% |
| No | 1377 | 48.9% |
| **Heart diseases** |  |  |
| Yes | 347 | 12.3% |
| No | 2471 | 87.7% |
| **Stroke** |  |  |
| Yes | 258 | 9.2% |
| No | 2560 | 91.8% |
| **MHD** |  |  |
| Yes | 545 | 19.3% |
| No | 2273 | 80.7% |

JHS: junior high school, SHS: senior high school

Table 2 identified variables that related to the MHD. Sex, residence type, educational level, employment status, obesity, hypertension, heart disease, stroke, and family history of MHD were significantly different between the MHD groups based on the Chi-square test; however, age, marital status, and duration of DM were comparable between the groups. The proportions of several parameters were significantly higher in the MHD group, i.e., female, rural residence, lower educational level, unemployed, obesity, hypertension, heart disease, family history of MHD, and stroke. These variables, as well as variables that were p ≤ 0.25, i.e., age category and marital status, then involved in the Binary logistic regression, and the final model of regression showed in Table 3.

**Table 2.** Subject characteristics based on MHD status

| Variables | MHD |  |  |  | p-value* | POR | % CI |
| --- | --- | --- | --- | --- | --- | --- | --- |
|  | Yes (n=545) |  | No(n=2273) |  |  |  |  |
|  | n | % | n | % |  |  |  |
| **Age category** |  |  |  |  | 0.11 | 1.17 | 0.968-1.407 |
| 60-69 years | 271 | 18.2 | 1218 | 81.8 |  |  |  |
| >69 years | 274 | 20.6 | 1055 | 79.4 |  |  |  |
| **Sex** |  |  |  |  | 0.001 | 1.56 | 1.282-1.901 |
| Female | 366 | 22.1 | 1289 | 77.9 |  |  |  |
| Male | 179 | 15.4 | 984 | 84.6 |  |  |  |
| **Residence** |  |  |  |  | 0.001 | 0.61 | 0.504-0.736 |
| Urban | 296 | 16.5 | 1503 | 83.5 |  |  |  |
| Rural | 249 | 24.4 | 770 | 75.6 |  |  |  |
| **Marital status** |  |  |  |  | 0.08 | 1.19 | 0.983-1.432 |
| Not married | 247 | 20.9 | 935 | 79.1 |  |  |  |
| Married | 298 | 18.2 | 1338 | 81.8 |  |  |  |
| **Educational level** |  |  |  |  | 0.001 | 2.49 | 1.932-3.200 |
| JHS or lower | 464 | 22.6 | 1585 | 77.4 |  |  |  |
| SHS or higher | 81 | 10.5 | 688 | 89.5 |  |  |  |
| **Employment status** |  |  |  |  | 0.003 | 1.35 | 1.111-1.647 |
| Unemployed | 364 | 21.1 | 1359 | 78.9 |  |  |  |
| Employed | 181 | 16.5 | 914 | 83.5 |  |  |  |
| **Obesity** |  |  |  |  | 0.001 | 5.51 | 4.072-7.468 |
| Yes | 100 | 52.9 | 89 | 47.1 |  |  |  |
| No | 445 | 16.9 | 2184 | 83.1 |  |  |  |
| **Hypertension** |  |  |  |  | 0.001 | 1.96 | 1.618-2.383 |
| Yes | 351 | 24.3 | 1090 | 75.6 |  |  |  |
| No | 194 | 14.0 | 1183 | 85.0 |  |  |  |
| **Heart diseases** |  |  |  |  | 0.02 | 1.37 | 1.047-1.785 |
| Yes | 83 | 23.9 | 264 | 76.1 |  |  |  |
| No | 462 | 18.7 | 2009 | 81.3 |  |  |  |
| **Stroke** |  |  |  |  | 0.001 | 2.29 | 1.733-3.022 |
| Yes | 86 | 33.3 | 172 | 66.4 |  |  |  |
| No | 495 | 19.0 | 2101 | 81.0 |  |  |  |
| **Family history of MHD** |  |  |  |  | 0.001 | 2.62 | 1.891-3.630 |
| Yes | 63 | 36.8 | 108 | 63.2 |  |  |  |
| No | 482 | 18.2 | 2165 | 81.8 |  |  |  |
| **Duration of DM** |  |  |  |  | 0.94 | 0.99 | 0.823-1.199 |
| ≤ 5 years | 245 | 19.4 | 1018 | 80.6 |  |  |  |
| ≥ 5 years | 300 | 19.3 | 1255 | 80.7 |  |  |  |

JHS: junior high school, SHS: senior high school, POR: prevalence odds ratio
\*Chi-square test

**Table 3.** Binary logistics regression of risk factors for MHD

| **Risk factors** | **p-value** | **POR** | **95%CI** |
| --- | --- | --- | --- |
| Family history of MHD | 0.001 | 2.43 | 1.707-3.471 |
| Female | 0.004 | 1.43 | 1.122-1.813 |
| Urban residence | 0.006 | 0.75 | 0.607-1.183 |
| Lower education | 0.001 | 1.93 | 1.464-2.533 |
| Obesity | 0.001 | 4.57 | 3.312-6.297 |
| Hipertension | 0.001 | 1.74 | 1.416-2.145 |
| Heart diseases | 0.006 | 1.49 | 1.123-1.973 |
| Stroke | 0.001 | 1.76 | 1.292-2.384 |

Nagelkerke R: 0.375; POR: prevalence odds ratio

Of the 11 variables involved in the Binary logistic regression model, eight parameters had statistically significance. Obesity (prevalence odds ratio [POR]=4.57; 95% CI: 3.312-6.297), family history of MHD (POR=2.43; 95% CI: 1.707-3.471), lower education (POR=1.93; 95% CI: 1.464-2.533), stroke (POR=1.76; 95% CI: 1.292-2.384), hypertension (POR=1.74; 95% CI: 1.416-2.145), heart disease (POR=1.49; 95% CI: 1.123-1.973), female (POR=1.43; 95% CI: 1.122-1.813), and urban residence (POR=0.75; 95% CI: 0.607-1.183) altogether were associated with MHD among elderly diabetics.

## Discussion

This cross-sectional study involved 2,818 elderly diabetics in Indonesia. Of them, 545 experienced MHD indicated that the prevalence of MHD among elderly diabetics in this population study was 19.3%. The current study updated the prevalence of MHD among elderly diabetics aged older than 60 years old, especially in Indonesia. A systematic review involved 248 studies and concluded a prevalence of 28% in people with type 2 diabetes experienced depression globally and 32% in Asia[8]. Diabetics aged older than 65 years old had a prevalence ratio of 21%[8], indicated similarly with the current study, while in the younger age (<65 years old) had a more prevalence ratio, i.e., 31%[8]. The female group had more prevalence than the male group, i.e., 34% and 24%, respectively[8]. Depression determination methods also influence the prevalence ratio; self-reported methods tend to had a higher prevalence (30%) than clinical diagnosis assessment (22%)[8]. The current study utilized self-reported methods using WHO-SRQ-20[22]; however, it found a lower number of prevalent compared to the previous review[8]. A previous systematic review of 26 studies involved all measurement assessments conducted in 2011 concluded that the prevalence of major depressive disorder in type 2 DM was 14.5% indicated lower number of prevalence ratio[23]. Another study observed diabetic patients in primary care aged more than 55 years, found the MHD prevalence of 19.1%[24].

Present study also found that the family history of MHD, sex, type of residence, educational level, obesity, hypertension, heart disease, and stroke altogether associated with MHD among elderly diabetics with the pseudo R (Nagelkerke) was 0.375. This finding explained that 37.5% of MHD determinants in this population study of elderly diabetics influenced by the mentioned factors, and the rest 62.5% explained by other factors that did not observe in the study. Present study involved many determinants; however, only those provided in RISKESDAS 2018 data. This study did not observe other pivotal determinants for MHD in diabetics. The determinants involved physical capability, insulin, and drug usage, ethnicity, detailed civil status (married, single, divorced, widowed), residence status (living alone, nuclear family, joint/extended living), family size, family income, pensioner status, smoking, alcohol, religion, glycaemic control, other sociodemographic and clinical health factors[8, 9, 11, 25–27]. The involved determinants in the study will potentially explain other parameters that influence MHD among elderly diabetics.

Interestingly, the current study found that obesity was associated with MHD in elderly diabetics with the highest POR, i.e., 4.57. A review study[28] concluded that obesity raises a 70% risk of MHD, while MHD increases the 40% risk of obesity. However, since this study did not observe longitudinally, so it cannot be determined whether obese occurred prior to MHD or vice versa. Another possibility in MHD prior to obesity, related to the use of its medication that one of the side effects is weight gain[29]. Future studies should be done to elucidate the relationship between obesity and MHD in diabetics, especially among the elderly involved, personality, cardiometabolic traits[30], and cognitive impairments[31].

Family history of MHD was found in this study as a risk factor with high POR, i.e., 2.43 means elderly diabetic with a family history of MHD had a risk 2.43 times compared to elderly diabetic without any family history. Indeed, in general, population, MHD is mostly influenced by the genetic and family factor[32]. Previous studies involved stress and epigenetics as a predictor of MHD in the general population[33]. Candidate genes were also studied and revealed that APOE, BDNF, SLC6A4 polymorphisms related to MHD in the general population[34]. Other studies revealed the contribution of inflammatory markers, such as interleukin (IL)-1β, IL-6, IL-10, monocyte chemoattractant protein-1, tumor necrosis factor-alpha, C-reactive protein, and phospholipase A2 contributed to the depression[35], that is also associated with type 2 diabetes[36].

Lower education level was another significantly associated factor with MHD in this study. Many other studies have reported a similar association of lower educational status with MHD in the diabetes population as well as in the general population[37–40]. Lower education level diabetics have limitations in coping[41] to deal with diabetes complications and other comorbidities as well as their general psychosocial problems. This study showed that elderly diabetics had almost two times the risk of experiencing MHD compared to higher education level. Present study determined lower education levels for elderly diabetics passed for junior high school (secondary education) or lower. Previous studies categorized level education in more detail, i.e., no schooling, primary education, secondary education, and tertiary education.[25].

This study also concluded that female elderly diabetics had a 43% higher risk of getting MHD compared to males. Either in the general population, diabetics, or chronic disease patients, females more likely to get MHD.[8, 42] Current study also found that elderly diabetics who lived in an urban area are less likely to experience MHD (POR=0.75) compared to diabetics live in a rural area. Most of the elderly diabetics in this study population were living in an urban area (63.8%); however, the proportion of MHD among elderly diabetics more likely occur in rural areas. Previous studies showed different conclusions regarding the higher prevalence of MHD in rural or urban areas[25, 40, 43], while other studies reported separately[11, 38, 44]. In developed countries, MHD tends to be higher in urban residents[45]. The current study also concluded a significant relationship between stroke, hypertension, and heart disease to MHD. The risk of stroke, hypertension, and heart disease is 76%, 74%, and 49% higher, respectively, compared to the absence of these conditions. Studies concluded that the presence of comorbidity more likely to experience MHD[11]. The more the number of comorbidities and additional illnesses, the higher the risk of getting MHD[11]. This condition also related to the physical limitations and capabilities, as well as the complicated clinical and health conditions involved drug usages[26].

## Conclusions

The prevalence of MHD among elderly diabetics in Indonesia was 19.3%. The risk factors for MHD among elderly diabetic subjects were family history of MHD, female, rural residence, low education, obesity, hypertension, heart disease, and stroke. The high prevalence of MHD among elderly diabetics, suggesting screening for psychological problems and educating elderly diabetic patients should be considered as routine components of diabetes care. Further studies should be conducted using clinical diagnosis assessment in a large population study, involved genetic factors, inflammatory markers, cardiometabolic traits, and other potential factors to elucidate the relationship between risk factors and the occurrence of MHD among elderly diabetic subjects as well as its mechanisms.

## Data Availability

The data used to support the findings of this study are available from the corresponding author Mahalul Azam upon request through the email address.

## Acknowledgments

We thank the Faculty of Sports Science, Universitas Negeri Semarang, for the Research Grant under the ID number 36.4.5/UN37/PPK.4.6/2020. We also thank the National Institute of Health Research and Development, Ministry of Health, the Republic of Indonesia, for providing RISKESDAS 2018 data.

## References

[1] International Diabetes Federation. IDF Diabetes atlas 9th edition. Brussel, Belgium, https://www.diabetesatlas.org (2019).

[2] International Diabetes Federation. IDF Clinical Practice Recommendations for managing Type 2 Diabetes in Primary Care International Diabetes Federation - 2017. 2017.

[3] Badan Penelitian dan Pengembangan Kesehatan. Laporan Nasional Riset Kesehatan Dasar: RISKESDAS (Indonesia Basic Health Survey) tahun 2018, http://labmandat.litbang.kemkes.go.id/images/download/laporan/RKD/2018/Laporan_Nasional_RKD2018_FINAL.pdf (2018).

[4] Badan Penelitian dan Pengembangan Kesehatan. Riset Kesehatan Dasar Riskesdas 2007. Jakarta, 2007.

[5] World Health Organization. Proportional mortality (% of total deaths, all ages). 2016.

[6] BPJS Kesehatan. Ringkasan Eksekutif Pengelolaan Program dan Laporan Keuangan Jaminan Sosial Kesehatan. 2016.

[7] Robinson DJ, Luthra M, Vallis M. Diabetes and Mental Health. Can J Diabetes 2013; 37: S87–S92.

[8] Khaledi M, Haghighatdoost F, Feizi A, et al. The prevalence of comorbid depression in patients with type 2 diabetes: an updated systematic review and meta-analysis on huge number of observational studies. Acta Diabetol 2019; 0: 20.

[9] Roy T, Lloyd CE. Epidemiology of depression and diabetes: A systematic review. J Affect Disord 2012; 142: S8–S21.

[10] Khan P, Qayyum N, Malik F, et al. Incidence of Anxiety and Depression Among Patients with Type 2 Diabetes and the Predicting Factors. Cureus; 11. Epub ahead of print 2019. DOI: 10.7759/cureus.4254.

[11] Sunny AK, Khanal VK, Sah RB, et al. Depression among people living with type 2 diabetes in an urbanizing community of Nepal. PLoS One 2019; 14: 1–11.

[12] De Sousa A, Lodha P. Geriatric Mental Heatlh: the challenges for India. J Geriatr Ment Heal 2018; 94–98.

[13] Bernstein CM, Stockwell MS, Gallagher MP, et al. Mental health issues in adolescents and young adults with type 1 diabetes: Prevalence and impact on glycemic control. Clin Pediatr (Phila) 2013; 52: 10–15.

[14] Sivertsen B, Petrie KJ, Wilhelmsen-Langeland A, et al. Mental health in adolescents with Type 1 diabetes: Results from a large population-based study. BMC Endocr Disord 2014; 14: 1–8.

[15] Eren I, Erdi Ö, Şahin M. The effect of depression on quality of life of patients with type II diabetes mellitus. Depress Anxiety 2008; 25: 98–106.

[16] Smith KJ, Pedneault M, Schmitz N. Investigation of anxiety and depression symptom co-morbidity in a community sample with type 2 diabetes: Associations with indicators of self-care. Can J Public Heal 2015; 106: e496–e501.

[17] Bruce DG, Davis WA, Dragovic M, et al. Comorbid Anxiety and Depression and Their Impact on Cardiovascular Disease in Type 2 Diabetes: The Fremantle Diabetes Study Phase II. Depress Anxiety 2016; 33: 960–966.

[18] Hendriks SM, Spijker J, Licht CMM, et al. Disability in anxiety disorders. J Affect Disord 2014; 166: 227–233.

[19] Naicker K, Johnson JA, Skogen JC, et al. Type 2 diabetes and comorbid symptoms of depression and anxiety: Longitudinal associations with mortality risk. Diabetes Care 2017; 40: 352–358.

[20] Yu M, Zhang X, Lu F, et al. Depression and Risk for Diabetes: A Meta-Analysis. Can J diabetes 2015; 39: 266–272.

[21] Tiki T. Prevalence and Associated Factors of Depression among Type 2 Diabetes Mellitus Patients on Follow up at Ambo General Hospital, Oromia Regional State, Ethiopia, Institutional Based Cross Sectional Study. J Depress Anxiety 2017; 06: 1–5.

[22] Netsereab TB, Kifle MM, Tesfagiorgis RB, et al. Validation of the WHO self-reporting questionnaire-20 (SRQ-20) item in primary health care settings in Eritrea. Int J Ment Health Syst 2018; 12: 61.

[23] Wang F, Wang S, Zong Q-Q, et al. Prevalence of comorbid major depressive disorder in Type 2 diabetes: a meta-analysis of comparative and epidemiological studies. Diabet Med 2019; 36: 961–969.

[24] McCombe G, Fogarty F, Swan D, et al. Identified mental disorders in older adults in primary care: A cross-sectional database study. Eur J Gen Pract 2018; 24: 84–91.

[25] Arambewela MH, Somasundaram NP, Ranjan Jayasekara HBP, et al. Prevalence of Depression and Associated Factors among Patients with Type 2 Diabetes Attending the Diabetic Clinic at a Tertiary Care Hospital in Sri Lanka: A Descriptive Study. Psychiatry J 2019; 2019: 1–8.

[26] Chen F, Wei G, Wang Y, et al. Risk factors for depression in elderly diabetic patients and the effect of metformin on the condition. BMC Public Health 2019; 19: 1063.

[27] Ganasegeran K, Renganathan P, Manaf RA, et al. Factors associated with anxiety and depression among type 2 diabetes outpatients in Malaysia: A descriptive cross-sectional single-centre study. BMJ Open 2014; 4: 1–8.

[28] Mannan M, Mamun A, Doi S, et al. Prospective Associations between Depression and Obesity for Adolescent Males and Females-A Systematic Review and Meta-Analysis of Longitudinal Studies. PLoS One 2016; 11: e0157240.

[29] Zimmermann U, Kraus T, Himmerich H, et al. Epidemiology, implications and mechanisms underlying drug-induced weight gain in psychiatric patients. J Psychiatr Res 2003; 37: 193–220.

[30] Franks PW, Atabaki-Pasdar N. Causal inference in obesity research. J Intern Med 2017; 281: 222–232.

[31] Brayne C, Gao L, Matthews F. Challenges in the epidemiological investigation of the relationships between physical activity, obesity, diabetes, dementia and depression. Neurobiol Aging 2005; 26 Suppl 1: 6–10.

[32] Köhler CA, Evangelou E, Stubbs B, et al. Mapping risk factors for depression across the lifespan: An umbrella review of evidence from meta-analyses and Mendelian randomization studies. J Psychiatr Res 2018; 103: 189–207.

[33] Park C, Rosenblat JD, Brietzke E, et al. Stress, epigenetics and depression: A systematic review. Neurosci Biobehav Rev 2019; 102: 139–152.

[34] Tsang RSM, Mather KA, Sachdev PS, et al. Systematic review and meta-analysis of genetic studies of late-life depression. Neurosci Biobehav Rev 2017; 75: 129–139.

[35] Barnes J, Mondelli V, Pariante CM. Genetic Contributions of Inflammation to Depression. Neuropsychopharmacol Off Publ Am Coll Neuropsychopharmacol 2017; 42: 81–98.

[36] Lontchi-Yimagou E, Sobngwi E, Matsha TE, et al. Diabetes mellitus and inflammation. Curr Diab Rep 2013; 13: 435–444.

[37] Sweileh WM, Abu-Hadeed HM, Al-Jabi SW, et al. Prevalence of depression among people with type 2 diabetes mellitus: A cross sectional study in Palestine. BMC Public Health; 14. Epub ahead of print 2014. DOI: 10.1186/1471-2458-14-163.

[38] Niraula K, Kohrt BA, Flora MS, et al. Prevalence of depression and associated risk factors among persons with type-2 diabetes mellitus without a prior psychiatric history: A cross-sectional study in clinical settings in urban Nepal. BMC Psychiatry; 13. Epub ahead of print 2013. DOI: 10.1186/1471-244X-13-309.

[39] Baumeister H, Härter M. Prevalence of mental disorders based on general population surveys. Soc Psychiatry Psychiatr Epidemiol 2007; 42: 537–546.

[40] Madkhali JM, Hakami AA, Dallak AH, et al. Prevalence and Associated Factors of Depression among Patients with Diabetes at Jazan Province, Saudi Arabia: A Cross-Sectional Study. Psychiatry J 2019; 2019: 1–6.

[41] Kara B, Açıkel CH. Predictors of coping in a group of Turkish patients with physical disability. J Clin Nurs 2012; 21: 983–993.

[42] Anandakumar D, Ratnatunga SS, Dayabandara M, et al. Depressive disorder in patients attending the outpatient department of a tertiary care hospital in Colombo. Ceylon Med J 2016; 61: 118–122.

[43] Li G, Mei J, You J, et al. Sociodemographic characteristics associated with adolescent depression in urban and rural areas of Hubei province: a cross-sectional analysis. BMC Psychiatry 2019; 19: 386.

[44] Asghar S, Hussain A, Ali SMK, et al. Prevalence of depression and diabetes: a population-based study from rural Bangladesh. Diabet Med 2007; 24: 872–877.

[45] Purtle J, Nelson KL, Yang Y, et al. Urban-Rural Differences in Older Adult Depression: A Systematic Review and Meta-analysis of Comparative Studies. Am J Prev Med 2019; 56: 603–613.

